# Deriving Imaging Biomarkers for Primary Central Nervous System Lymphoma Using Deep Learning

**DOI:** 10.1101/2024.09.16.24313435

**Authors:** Joshua Zhu, Michela Destito, Chitanya Dhanireddy, Tommy Hager, Sajid Hossain, Saahil Chadha, Durga Sritharan, Anish Dhawan, Keervani Kandala, Christian Pedersen, Nicoletta Anzalone, Teresa Calimeri, Elena De Momi, Maria Francesca Spadea, Mariam S. Aboian, Sanjay Aneja

## Abstract

**Purpose:** Primary central nervous system lymphoma (PCNSL) is typically treated with chemotherapy, steroids, and/or whole brain radiotherapy (WBRT). Identifying which patients benefit from WBRT following chemotherapy, and which patients can be adequately treated with chemotherapy alone remains a persistent clinical challenge. Although WBRT is associated with improved outcomes, it also carries a risk of neuro-cognitive side effects. This study aims to refine patient phenotyping for PCNSL by leveraging deep learning (DL) extracted imaging biomarkers to enable personalized therapy.

**Methods:** Our study included 71 patients treated at our institution between 2009-2021. The primary outcome of interest was overall survival (OS) assessed at one-year, two-year, and median cohort survival cutoffs. The DL model leveraged an 8-layer 2D convolutional neural network which analyzed individual slices of post-contrast T1-weighted pre-treatment MRI scans. Survival predictions were made using a weighted voting system related to tumor size. Model performance was assessed with accuracy, sensitivity, specificity, and F1 scores. Time-dependent AUCs were calculated and C-statistics were computed to summarize the results. Kaplan-Meier (KM) survival analysis assessed differences between low and high-risk groups and statistically evaluated using the log-rank test. External validation of our model was performed with a cohort of 40 patients from an external institution.

**Results:** The cohort’s average age was 65.6 years with an average OS of 2.80 years. The one-year, two-year, and median OS models achieved AUCs of 0.73 (95% C.I., 0.60-0.85), 0.70 (95% C.I., 0.58-0.82), and 0.73 (95% C.I., 0.58-0.82) respectively. KM survival curves showcased discrimination between low and high-risk groups in all models. External validation with our one-year model achieved AUC of 0.64 (95% C.I., 0.63-0.65) and significant risk discrimination. A sub-analysis showcased stable model performance across different tumor volumes and focality.

**Conclusions:** DL classifiers of PCNSL MRIs can stratify patient phenotypes beyond traditional risk paradigms. Given dissensus surrounding PCNSL treatment, DL can augment risk stratification and treatment personalization, especially with regards to WBRT decision making.

## 1 Introduction

Primary central nervous system lymphoma (PCNSL) is a rare, aggressive form of extranodal non-Hodgkin lymphoma limited to the central nervous system (CNS), involving the brain, cerebrospinal fluid, and eyes without evidence of systemic spread [1]. Treatment for PCNSL has evolved over the years, and currently remains an active area of research.

The current PCNSL treatment paradigm consists of an induction and consolidation phase [2, 3]. The induction phase typically consists of a high-dose methotrexate (MTX) based chemotherapy. For patients with favorable disease response to induction chemotherapy, a decision is made whether to proceed with the consolidation phase, which incorporates whole-brain radiotherapy (WBRT) [4].

Although WBRT is associated with improved outcomes, WBRT has been contentious due to risk of neuro-cognitive side effects [5, 6]. Recent clinical trials have scrutinized the necessity and efficacy of WBRT in the PCNSL treatment paradigm. One trial investigating a chemotherapy-only approach (i.e. without WBRT) found that overall survival (OS) was not compromised, while progression free survival (PFS) was reduced [7]. However, due to trial limitations and conflicting evidence from subsequent studies, it remains unclear whether an intensive chemotherapy-only regimen can truly replace WBRT in terms of disease control [8, 9, 10], . More recently, trials have investigated the efficacy of reduced dose WBRT as a consolidation treatment, with preliminary findings indicating that disease control is comparable to full dose WBRT, while substantially decreasing neurotoxicity rates [11, 12].

Alongside an evolving treatment landscape, the management of PCNSL is further complicated by heterogeneities within the patient population. Diverse patient characteristics, which correlate with treatment response, underscores the importance and challenge of identifying high-risk patient phenotypes early in the disease course [13]. Currently, clinicians use PCNSL prognostic indices, most commonly the International Extranodal Lymphoma Study Group (IELSG) and Memorial Sloan Kettering Cancer Center (MSKCC) scales, to determine the aggressiveness of a patient’s cancer and guide clinical management [14, 15]. These scales account for prognostic factors, such as patient demographics, laboratory values, Karnofsky Performance Status, and tumor characteristics, to stratify the patient into a three-group system (low-, intermediate-, and high-risk). However, although these indices assist in general clinical guidance, they do not reflect more recent progress in PCNSL treatment, such as WBRT protocols and rituximab availability [16].

Deep learning (DL) has emerged as a promising technology to assist in PCNSL patient risk stratification. DL is a class of machine learning which uses artificial neural networks to automatically extract large amounts of quantitative features from imaging data, with minimal prior medical knowledge and feature selection. Related PCNSL work has focused on using DL to automatically segment tumors and discriminate between PCNSL and glioblastoma, with most studies reporting excellent performance [17, 18, 19, 20, 21]. Apart from this application, there has been limited research that leverages DL to predict clinical outcomes for PCNSL patients [22].

Our study aims to build a DL model which extracts imaging biomarkers that aid in PCNSL patient phenotyping to enable personalized therapy, i.e. whether patients will benefit from WBRT. Our work extends the previous lines of research from both data stream and model architecture perspectives. This study will be the first to utilize segmented T1-weighted magnetic resonance images (MRI) as data inputs in a DL model predicting patient OS. We also develop a novel convolutional neural network (CNN) architecture that is trained to recognize important tumor biomarkers through a segmentation task, then leverages learned imaging features to make binary OS predictions. By using this model to stratify patients into low and high-risk phenotypes, we demonstrate how MRI-derived biomarkers can guide the personalization of chemoradiation treatments in PCNSL patients.

## 2 Methods

### 2.1 Dataset

This study was approved by the Institutional Review Board (IRB) and was compliant with the Health Insurance Portability and Accountability Act (HIPAA). De-identified data was used, and no protected health information was needed. Clinical characteristics for all patients was collected by way of retrospective chart review and informed consent was waived due to the retrospective nature of this study.

The institutional database at Yale New Haven Hospital was queried for patients with a diagnosis for PCNSL from January 2009 until October 2021.The minimum imaging requirements included a pre-operative MRI study before histopathological diagnosis, using a gadolinium T1-weighted sequence. All selected studies were obtained using clinical 1.5 or 3 Tesla scanners. Patients with disease involvement outside of the CNS were excluded. A comprehensive flowchart of the number of patients included in each analysis, along with a description of exclusion criteria, is provided in **Supplemental Figure S1**. The final study cohort included 71 patients.

For all patients in the study cohort, clinical data regarding age, gender, immune status, and OS were recorded. The primary outcome of interest in this study is OS, which calculated with three separate cutoff points: one year, two years, or median survival of the cohort, which was 677 days. Positive and negative OS cases were assigned with survival greater and less than the cutoff point respectively.

For our external validation dataset, an institutional database at the San Raffaele Hospital in Milan, Italy was queried for patients with a diagnosis for PCNSL from January 2009 until October 2021. The minimum imaging requirements included a pre-operative MRI study before histopathological diagnosis, using a gadolinium T1-weighted sequence. Selected studies were obtained using different scanners, including Philips Medical System Achieva and Whole, GE Medical System/Optima MR450, and SIEMENS AERA. The final external validation cohort included 40 patients.

### 2.2 Image Preprocessing

Image preprocessing was performed with the following standard techniques — (1) skull stripping using the HD-BET prediction algorithm (2) image corrections for B1-field variations using the N4 bias-field correction algorithm (3) intensity normalization with histogram matching based on landmarks learned on a subset of images as proposed by Nyúl and Udupa et al. (1999) and (4) uniform voxel resampling to the median voxel size determined across the dataset [23, 24, 25]. To enhance the model’s focus on relevant tumor biomarkers and reduce extraneous noise, non-tumor slices were removed from the whole brain images. Finally, an extensive data augmentation was performed on preprocessed images for the train set including random rotation, horizontal flipping, and gaussian blurring. Segmentation of brain tumors was performed by board-certified neuroradiologists (C.P. and M.A.). These served as ground truth labels for the model during segmentation training. **Figure 1** provides an overview of the project workflow and image preprocessing pipeline.

**Figure 1:**
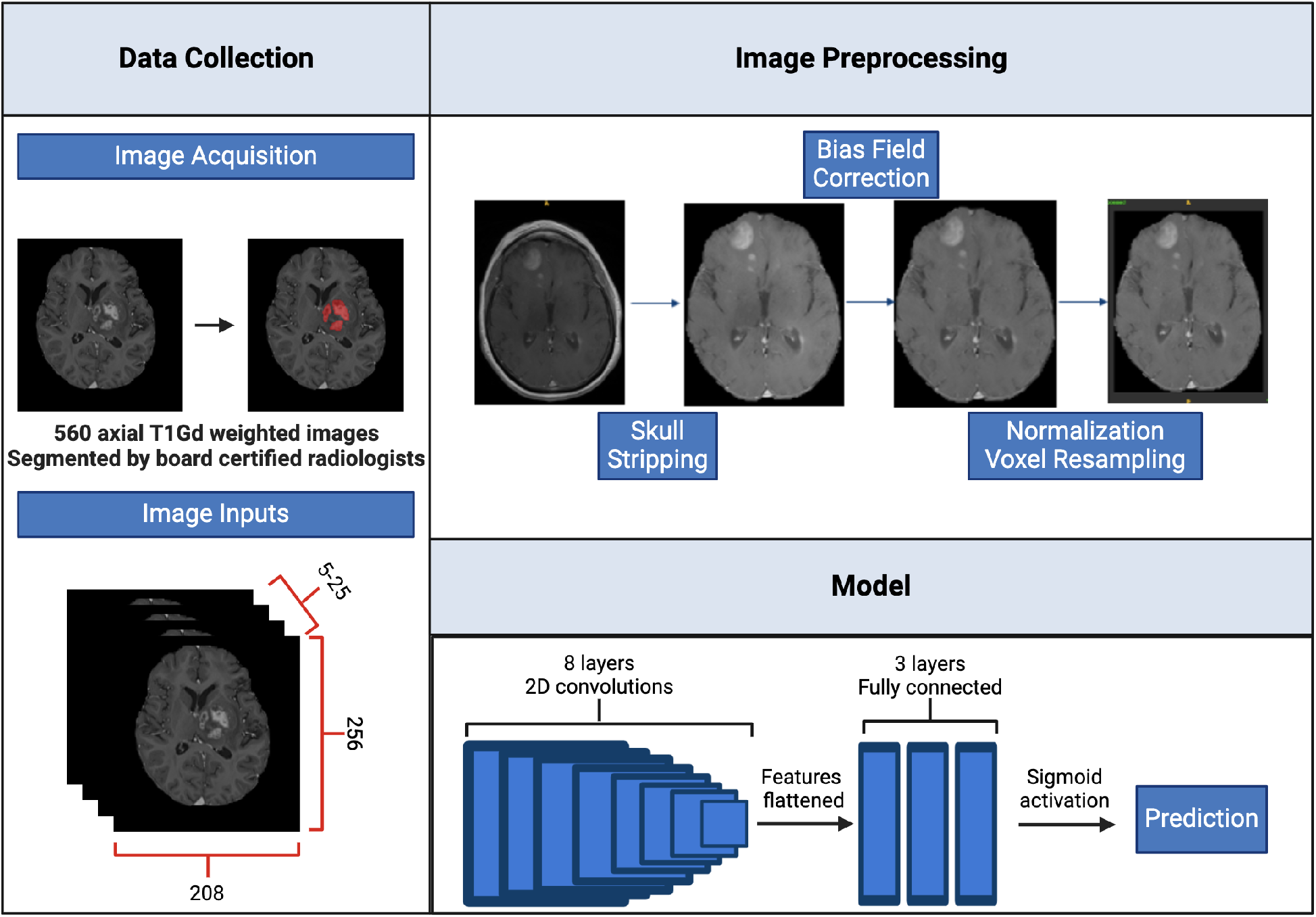
Overview of overall project workflow and image preprocessing pipeline.

### 2.3 Model Architecture and Training

The CNN employed in this study was trained in two phases: segmentation and classification.

During the segmentation phase, an unmodified UNet, as described by Ronneberger et al. (2015), was created for a tumor segmentation task [26]. The architecture included an 8-layer CNN architecture, with a contracting path capturing image context through successive convolutional and max-pooling layers, and an expansive path for precise localization using up-convolutional layers and skip connections. Regularization was applied via dropout layers with 50% probability, and training images included data augmentations. The model employed a hybrid loss function combining Dice Loss and Binary Cross-Entropy with Logits Loss, optimized using Stochastic Gradient Descent with an initial learning rate (lr) of 1e-3, a mini-batch size of 16 2D slices, and a weight decay of 1e-3.

To enhance feature recognition, we pretrained the UNet model for its segmentation task on a glioblastoma dataset from the same institutional database. The pretraining dataset comprised of gadolinium T1-weighted sequences from 418 glioblastoma patients [27]. MRI scans were obtained from the same institution, tumor segmentations were performed by the same board-certified neuroradiologists, and image preprocessing followed the same pipeline as the primary PCNSL dataset. Following pretraining, we applied transfer learning to adapt and finetune the model to the primary PCNSL dataset.

In the subsequent classification phase, the trained UNet model was repurposed for OS survival analysis. The contracting path of the UNet, which encapsulates information about tumor morphology and aggressiveness, was frozen and fed into a new fully connected network. Namely, a flattening operation was applied to the last convolutional layer to extract flattened features from each image. These features were inputted into three fully connected layers with LeakyReLU activation and dropout for regularization, culminating in a sigmoid activation to output the OS prediction [28]. The prediction of the network was based on an individual slice of a patient’s MRI (“slice-level prediction”).

To extend the predictions from individual MRI slices (“slice-level prediction”) to a patient’s overall prognosis, a voting system was employed to make an aggregate prediction (“patient-level prediction”). Our voting system took all slice-level predictions for a given patient, and added a weight based on the size of the tumor in the prediction image. This aggregate weighted sum was assessed against a calibrated threshold to determine the final patient-level prediction.

The equation for patient-level prediction from MRI slices is described as

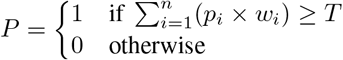

where *P* is the patient level prediction, *p*_*i*_ represents the binary prediction for each slice *i, w*_*i*_ denotes the weight based on tumor size in slice *i*. The sum aggregates predictions across all *n* slices, with *P* set to 1 if the sum is greater than or equal to *T* (threshold), indicating tumor presence, otherwise 0.

### 2.4 Evaluation metrics

Patient records included both categorical and continuous clinical data elements. Categorical variables were analyzed using frequencies, comparing groups using chi-squared or Fisher Exact tests. Continuous variables were presented as mean ± standard deviation, comparing groups using Wilcoxon rank sum tests.

Model performance during the segmentation training and finetuning phases was assessed via Dice similarity coefficient on the validation set [29]. Model performance during the classification phase was assessed on internal and external validation datasets via accuracy, sensitivity, specificity, F1-score, and the area under the receiver operating characteristic curve (AUC). Receiver Operating Characteristic (ROC) curves were generated using the logits produced by each model to assess model performance across different threshold values. Confidence intervals (C.I.) were calculated using 200 randomly generated bootstrapped samples, with replacement, from the original data set to create samples of the same size as the original. The 2.5^th^ and 97.5^th^ percentiles of the bootstrap distributions were then used to define the 95% confidence intervals.

Clinical viability of the model performance was assessed with Kaplan-Meier (KM) Survival Analysis and time-dependent AUCs. Patients were categorized as low or high risk based on model prediction of survival or death at one year respectively. Differences between risk groups were statistically evaluated using the log-rank test. Time-dependent AUCs were calculated at various time points to measure the discriminatory ability of each model over time, and C-statistics were computed to summarize the AUC results.

A sub-analysis of model performance was conducted, focusing on categorizations based on tumor volume and morphology. The median tumor volume for the cohort was 10,319 mm^3^, with tumors at or below designated as “below 50^th^ percentile volume,” while those above were termed “above 50^th^ percentile volume.” Multifocal tumors were defined as those presenting with multiple distinct regions of interest, and the remaining tumors were defined as unifocal.

### 2.5 Interpretability Methods

For each patient, the MRI slice with the largest tumor volume, as assessed by the overall size of segmentation mask, was selected. From this key slice, we extracted a single vector of deeply learned features by flattening the output feature maps from the CNN’s final convolutional layer. Each of the unique features was normalized to a scale of 0 to 1 by subtracting the minimum value and dividing by the maximum value across each cohort. To explore the patterns within these features, unsupervised hierarchal clustering of these normalized features was performed on the training set. The clusters identified were then applied to both internal and external validation sets to verify the reproducibility and relevance of these features across different data sets.

## 3 Results

### 3.1 Patients

From January 2009 to October 2021, a total of 71 patients with biopsy proven PCNSL were treated at our institution (**Table 1**). There were 35 male and 36 female patients included in this study. The average age was 65.6 years with an average OS of 2.80 years. At least 19 patients were immunocompromised at the time of diagnosis, including four patients who were HIV positive, six patients with a history of prior solid organ transplant, five patients with a history of an autoimmune disease, and 14 patients on immunosuppressive therapy at the time of diagnosis.

**Table 1:** Baseline demographics and disease characteristics of training and validation cohorts. P-value comparisons are conducted between the training and internal validation cohorts, as well as between the training and external validation cohorts.

| Characteristics | Overall (n = 71) | Training (n = 57) | Internal Validation (n = 14) | p-value | External Validation (n = 40) | p-value |
| --- | --- | --- | --- | --- | --- | --- |
| **Age (years)** | 65.6 ± 14.6 | 65.3 ± 14.6 | 67.5 ± 14.22 | 0.069 | 68.8 ± 10.9 | 0.18 |
| **Sex** |  |  |  | 0.56 |  | 0.05 |
| Male | 35 (49.3%) | 27 (47.4%) | 8 (57.1%) |  | 12 (30.0%) |  |
| Female | 36 (50.7%) | 30 (52.6%) | 6 (42.9%) |  | 28 (70.0%) |  |
| **Overall survival (days)** | 1021.5 ± 1054.8 | 982.8 ± 992.4 | 1179.3 ± 1309.9 | 0.44 | 1016.3 ± 1146.6 | 0.74 |
| < 1 year | 30 (42.3%) | 23 (40.4%) | 7 (50.0%) |  | 21 (52.5%) |  |
| > 1 year | 31 (43.7%) | 24 (42.1%) | 7 (50.0%) |  | 19 (47.5%) |  |
| **Tumor Multiplicity** |  |  |  | 0.77 |  | 0.47 |
| Unifocal | 34 (47.9%) | 28 (49.1%) | 6 (42.9%) |  | 22 (55.0%) |  |
| Multifocal | 37 (52.1%) | 29 (50.9%) | 8 (57.1%) |  | 18 (45.0%) |  |
| **ASCT** |  |  |  | 0.27 |  | 0.96 |
| Yes | 15 (21.1%) | 14 (24.6%) | 1 (7.14%) |  | 9 (22.5%) |  |
| No | 53 (74.6%) | 41 (71.9%) | 12 (85.7%) |  | 31 (77.5%) |  |
| Unknown | 3 (4.22%) | 2 (3.51%) | 1 (7.14%) |  | 0 |  |
| **Immune Status** |  |  |  | 0.18 |  | <0.01 |
| Immunocompetent | 52 (73.2%) | 44 (77.2%) | 8 (57.1%) |  | 40 (100%) |  |
| Immunosuppressed | 19 (26.8%) | 13 (22.8%) | 6 (42.9%) |  | 0 (0%) |  |
| **Proportion Necrosis** |  |  |  | 0.42 |  | — |
| 0% | 4 (5.63%) | 3 (5.26%) | 1 (7.14%) |  | — |  |
| <5% | 43 (60.56%) | 33 (57.9%) | 10 (71.4%) |  | — |  |
| 6-33% | 17 (23.94%) | 16 (28.1%) | 1 (7.14%) |  | — |  |
| 34-67% | 5 (7.04%) | 3 (5.26%) | 2 (14.3%) |  | — |  |
| 68-95% | 2 (2.81%) | 2 (3.51%) | 0 (0.00%) |  | — |  |
| >95% | 0 (0.00%) | 0 (0.00%) | 0 (0.00%) |  | — |  |

From January 2010 to November 2019, a total of 40 patients with biopsy proven PCNSL were treated at San Raffaele Hospital in Italy. This dataset included only immunocompetent patients, and significant differences with the internal cohort was observed in patient sex. There were no significant differences observed in other clinical characteristics.

### 3.2 Segmentation Phase

Our CNN was pretrained on our glioblastoma dataset for over 1000 epochs and in one validation fold split. The pretrained model achieved a Dice coefficient of 0.94 on the training cohort and 0.80 on the validation cohort. This model was finetuned on the study PCNSL dataset for over 200 epochs and in one validation fold split. The finetuned model achieved a Dice achieved of 0.92 on the training cohort and 0.83 on the validation cohort.

### 3.3 Binary Classification Phase

After adapting the CNN to the OS binary classification task as described, three separate models were developed for the one year, two year, and median OS cutoffs. Each model was trained for over 100 epochs and in one validation fold split. Slice-level accuracy ranged between 0.69-0.71. After implementing the voting system, the final patient-level accuracy improved to 0.70-0.73 (**Supplemental Table S1**,**S2**).

We use our best performing model, the one-year OS model, as a representative model for all subsequent main analysis. Our model achieved an AUC and accuracy of 0.75 and 0.70 on the internal validation cohort, and 0.64 and 0.67 on the external validation cohort respectively (**Figure 2**). KM curves, assessing clinical viability, demonstrated discrimination between low and high risk groups in both the internal (p = 0.025) and external (p = 0.002) validation cohorts (**Figure 3**). Comprehensive results for the two-year and median OS models are found in our supplement.

**Figure 2:**
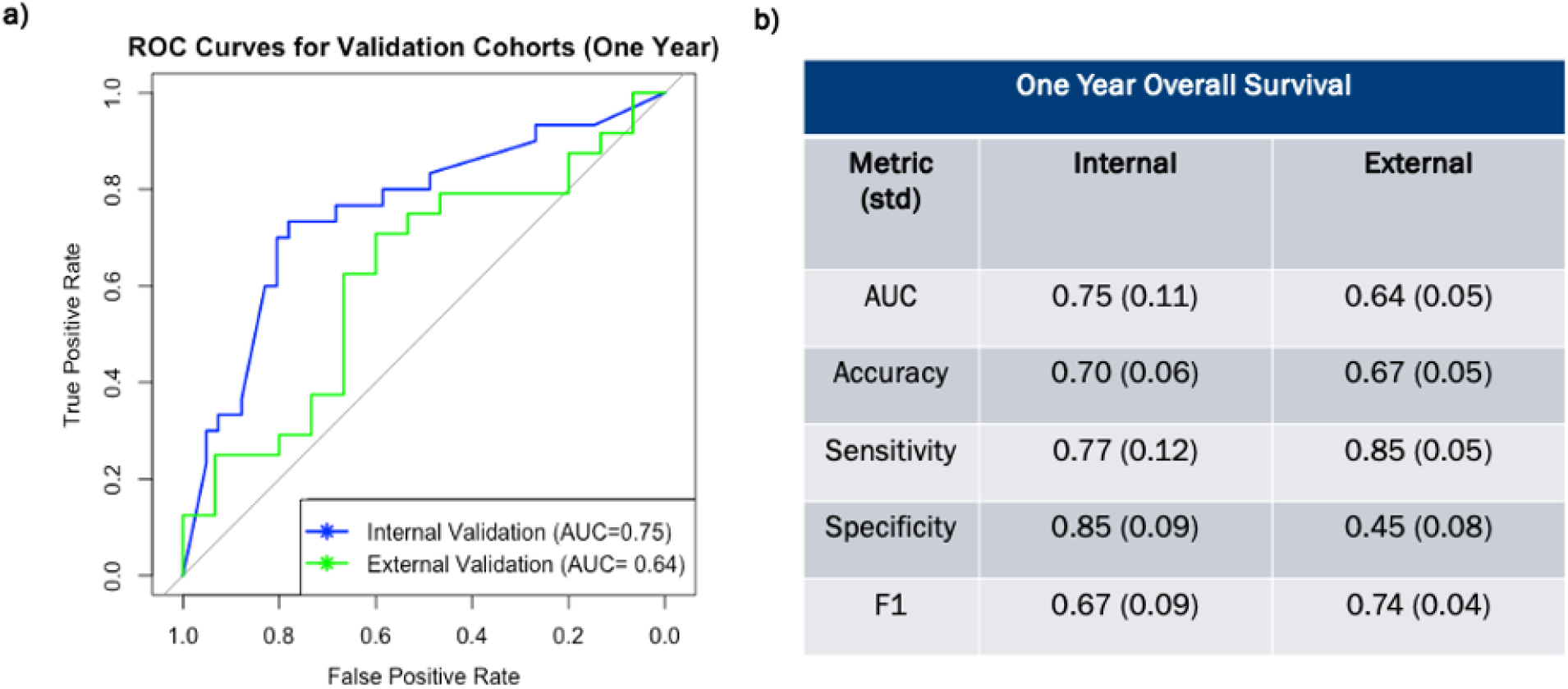
**a)** Receiver Operating Characteristic curves for one year survival model on the internal and external validation cohorts. Curve were generated using model logits, i.e. the raw prediction scores before binary prediction, to assess continuous prediction scores. The AUC values indicate the overall performance of each model, with higher AUC values representing better discriminative ability. **b)** Model evaluation metric across internal and external validation cohorts.

**Figure 3:**
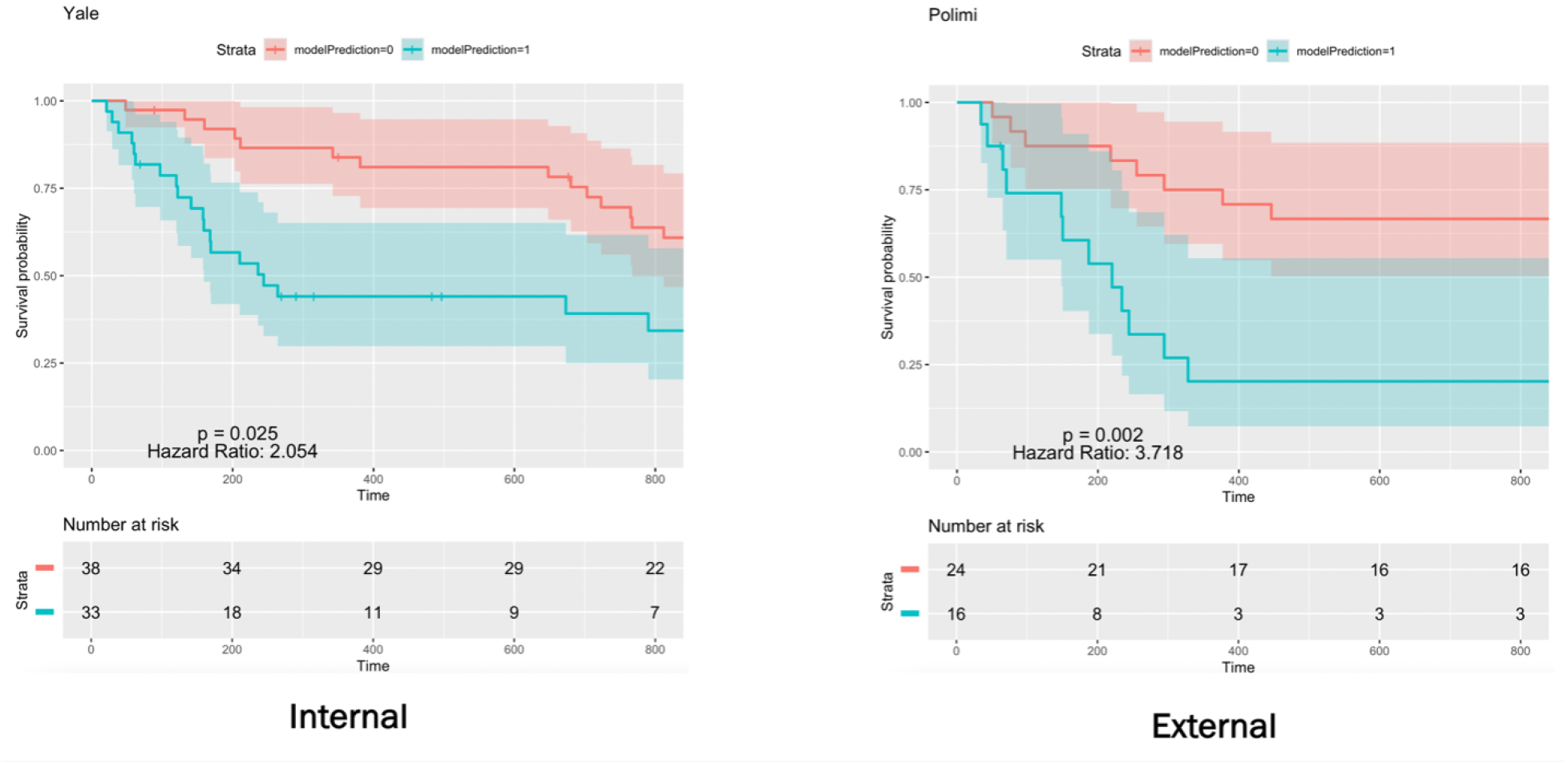
Kaplan-Meier survival curves comparing the performance of one year overall survival model across internal and external validation cohort. Each curve represents the survival probability over time for the low-risk group (red line) and high-risk group (blue line). The log-rank test was used to assess statistical differences between the models, with p-value, C-statistic, and Hazard ratio provided in the plot.

### 3.4 Subgroup Analysis

Model performance remained consistent in subgroup analysis. ROC curves showed performance improvement within the above 50^th^ percentile volume and multifocal cohorts compared to below 50^th^ percentile volume and unifocal cohorts. KM curves demonstrated an overall ability for the model to discriminate between risk groups (**Figure 4**).

**Figure 4:**
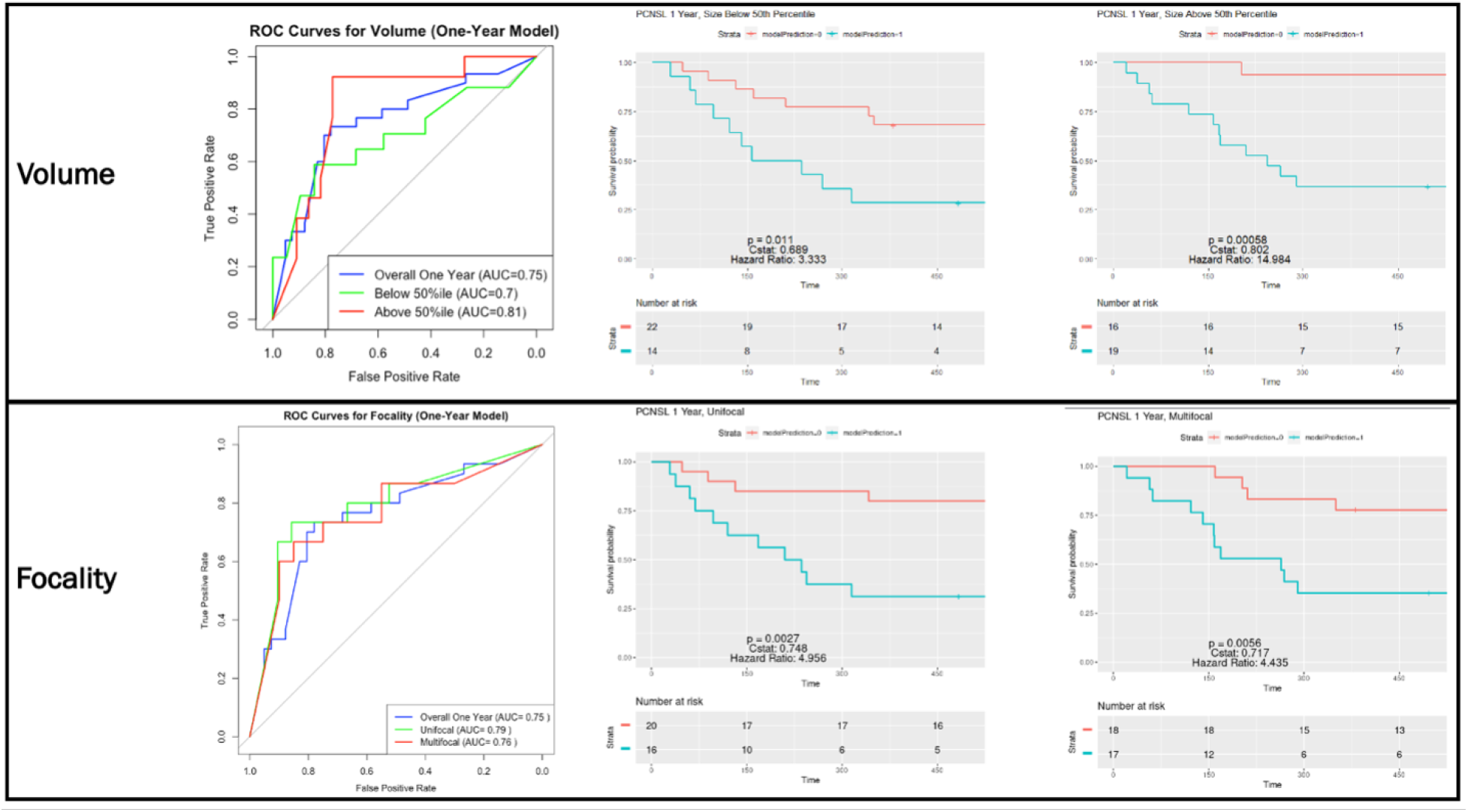
Validation of bioimaging marker across tumor size and multiplicity using one year overall survival model. Volume ROC curves were generated with the model performance at baseline (blue line), with below 50^th^ percentile volume cohort (green line), and with above 50^th^ percentile volume cohort (red line), where the 50^th^ percentile volume of the overall cohort was 10,319 mm^3^. Focality ROC curves were generated with the model performance at baseline (blue line), with unifocal cohort (green line), and multifocal cohort (red line). Kaplan Meier survival curves were generated to assess the performance of the model across cohorts.

### 3.5 Interpretability

We used an unsupervised hierarchal clustering method on deeply learned features extracted from the flattening of the last layer our CNN. For hierarchal clustering of features, two distinct phenotypic patterns are observed across training and validations sets corresponding to OS predictions (**Figure 5**). Across the training and validation sets, some overlap in the phenotypic patterns was observed, suggesting that the model is successfully extracting features related to OS.

**Figure 5:**
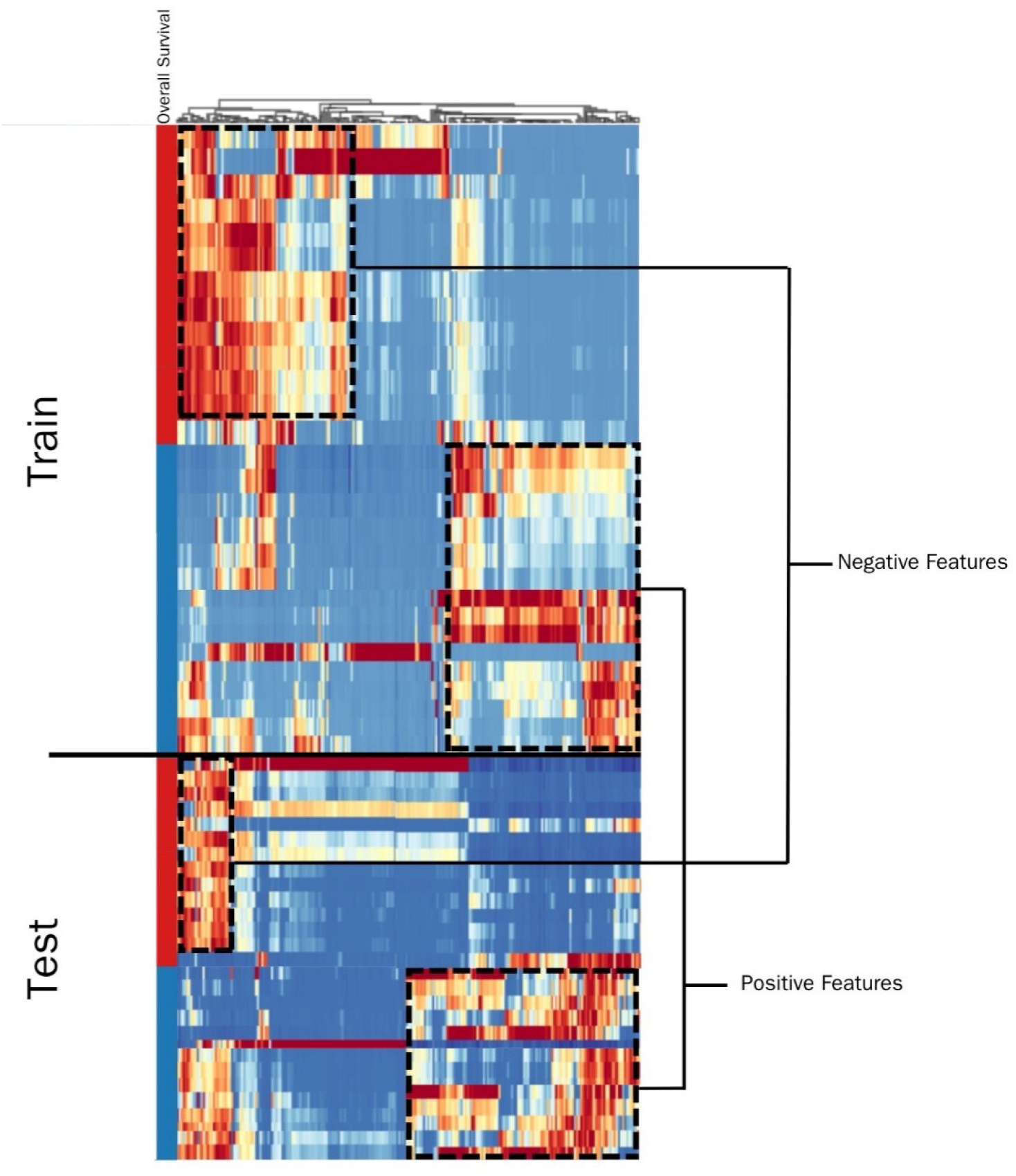
Interpretability of bioimaging marker with unsupervised clustering of the deeply learned features output by the last layer of the one year OS model. There are two distinct biomarker patterns representing OS less than one year (red) and greater than one year (blue).

## 4 Discussion

In clinical practice, PCNSL poses complex treatment decision-making challenges. To this end, we developed a novel DL approach to aid in risk stratification of patients based on OS. Our approach is the first to leverage MRI-derived imaging biomarkers extracted during PCNSL segmentation as prognostic indicators. This work can guide future personalization of chemoradiation treatment.

In this study, we developed a novel UNet-based CNN that extracted imaging biomarkers to effectively stratify patients into distinct risk categories. Our voting system further improved model accuracy to clinically significant values across all validation cohorts. KM survival analysis demonstrated our model’s ability to discriminate between risk groups. The successful implementation of our DL approach showcases that MRI imaging features are not only a quantifiable data stream, but also meaningful in clinical contexts. Our model was able to separate patients based on OS predictions, identifying low-risk patients who may be candidates for de-escalation after induction chemotherapy, and high-risk patients who may benefit more from WBRT. This patient discrimination helps facilitate decision-making in clinical practice, particularly in personalizing treatment plans to optimize patient outcomes.

We also demonstrate our model’s reliability through testing on an external dataset, and tumor subtypes by volume and focality. External validation on the San Raffaele Hospital dataset showcases our model’s robustness, especially within an Italian population with different patient characteristics and treatment protocols. Subgroup analysis showcases the overall stable performance of our model in all groups apart from the median OS model for below 50th percentile volume cohort. Notably, ROC curve analysis showed a marginal improvement in performance with larger and multifocal tumors. This is likely due to a variety of factors, including improved signal in larger tumor regions, and intrinsic challenges related to the CNN’s ability to effectively capture subtle features of smaller tumors in its filters. Additionally, pooling layers can cause the loss of fine details that are critical for identifying small tumors. Overall, the ability for our model to predict OS across tumor subtypes is particularly important as tumor morphology is indicative of varying clinical challenges and prognoses [30]. Our model’s ability to decode different tumor morphologies is critical in formulating personalized treatment plans tailored to specific patient characteristics and risks.

From a network perspective, our study pioneers a novel approach employing a CNN architecture that extracts 2D features, where early layers focus on capturing fundamental image characteristics such as edges and textures. Our results indicate that these basic elements extracted from medical imaging are not merely structural details, but can serve as critical biomarkers that are indicative of disease prognosis. These findings align with previous research, which has linked volumetric features, such as VASARI descriptors, to clinical outcomes [31, 32]. Moreoever, the 2D features extracted by our CNN can be seen as effective proxies for the more complex 3D patterns that describe a tumor’s structure and pathology. Thus, our study also suggests a pathway in reducing computational demands of future model development for clinical applications by utilizing the simpler 2D features from medical imaging.

Although the main goal of this study was prediction of clinical outcomes, we presented several innovative approaches the tumor segmentation portion of our experiment. Firstly, we opted to pretrain on a glioblastoma data. Because PCNSL and glioblastoma show similarities in their features, such as contrast-enhancing tumor, potential central necrosis, FLAIR-hyperintense tumor parts, and surrounding edema, we hypothesized that features useful for the detection and segmentation of glioblastoma might also be applied to PCNSL [33, 34, 35] [32-34]. In the finetuning phase, we hypothesized that the model would discern differences between PCNSL, which usually consists of several scattered homogenous enhanced tumor parts, versus glioblastoma, which usually consists of a single tumor with a ringlike zone of contrast-enhancement surrounding necrosis and intralesional hemorrhage.

To our knowledge, there is only one other study that has developed a DL model for segmentation of PCNSL. Pennig et al. (2021) leveraged a DL model developed originally for glioblastoma segmentation, and evaluated its efficacy on a PCNSL dataset [36]. This study reported a Dice coefficient of 0.80 for total tumor volume and 0.74 for core in pretreatment scans. Our methodology extends the current research by adding a finetuning component, and reports an improvement in segmentation efficacy with a Dice score of 0.82 for total tumor volume.

Interpretability issues remain a major barrier to translating recent breakthroughs in imaging biomarkers from bench to clinic. To this end, we presented a qualitatively interpretable likelihood scale to promote the interpretability of our imaging biomarkers. Our results show a concordance between regions on MRI imaging corresponding to high-risk PCNSL phenotypes and regions recognized by our deep learning models to inform OS classification predictions. We hope that these tools can contribute to a shared decision-making process, which is especially relevant in the oncological setting, in which prognostic biomarkers must be plainly explainable to patients.

Currently, treating clinicians often encounter challenges in recommending PCNSL treatments when they are inadequately informed about a patient’s prognosis. The difficulty in characterizing tumors, wide-ranging aggressiveness, and often unpredictable patient response to chemotherapeutic options adds more complexity to the clinical decision-making process. Traditional clinicopathologic paradigms of risk stratification, such as IELSG and MSKCC, have limited prognostic utility as they predate the advent of modern, intensive treatment protocols [37, 38]. Recent efforts to update these PCNSL indices have involved incorporation of clinical and molecular markers to provide personalized data [39, 40, 41]. Unfortunately, these approaches have largely been limited by small sample sizes and manual feature extraction, which is dependent on manual tumor delineation by experts. Incorporating a DL-based model, such as the one developed in this study, addresses these limitations by automating feature extraction and improving the accuracy of patient classification and prognosis prediction.

We recognize that our study has several limitations to be addressed. Firstly, the retrospective nature of the study and patient cohort size limits the generalizability of our results. More evidence utilizing external validation datasets from multiple centers is needed, in addition to prospective clinical trial validation before application into the clinic. Additionally, we acknowledge that our interpretability scale is prone to incorrect interpretation of the original reader. As interpretable AI methodologies develop, future studies could utilize more robust techniques that visualize the patterns of our model. Finally, during the course of this study, we experimented with several voting systems. Although we found success with the weighted voting system, future research might experiment with different methodologies.

## Data Availability

All data produced in the present study are available upon reasonable request to the authors.

## A Supplementary Data

**Table S1:** Slice-level model performance evaluation across the overall cohort.

| Metric | One Year | Two Year | Median |
| --- | --- | --- | --- |
| **AUC (std)** | 0.74(0.11) | 0.75(0.15) | 0.73(0.09) |
| **Accuracy (std)** | 0.70(0.06) | 0.69(0.06) | 0.71(0.05) |
| **Sensitivity (std)** | 0.77(0.12) | 0.78(0.10) | 0.71(0.07) |
| **Specificity (std)** | 0.85(0.09) | 0.59(0.11) | 0.68(0.07) |
| **F1 (std)** | 0.67(0.09) | 0.74(0.03) | 0.73(0.05) |

**Table S2:**
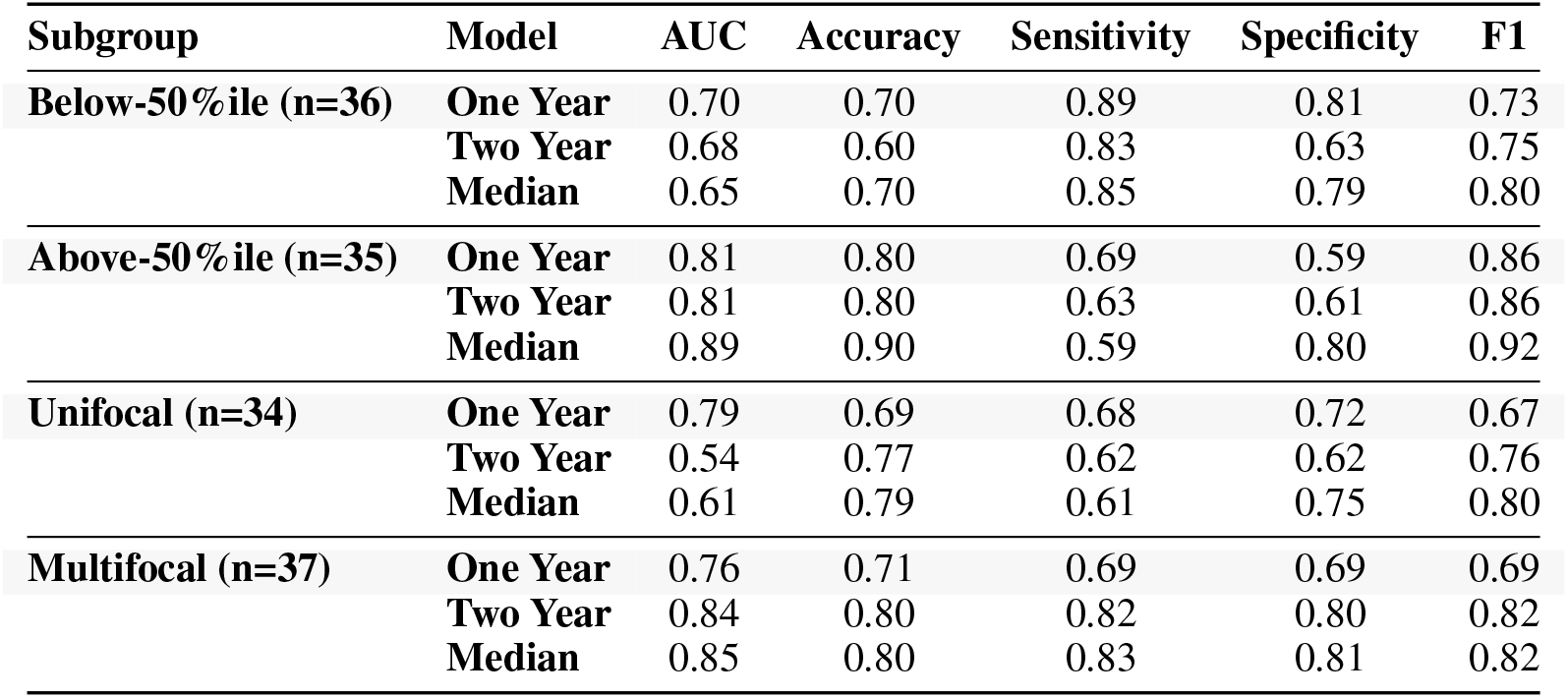
Sub-analysis of tumor size and multiplicity on model performance.

| Subgroup | Model | AUC | Accuracy | Sensitivity | Specificity | F1 |
| --- | --- | --- | --- | --- | --- | --- |
| **Below-50%ile (n=36)** | **One Year** | 0.70 | 0.70 | 0.89 | 0.81 | 0.73 |
|  | **Two Year** | 0.68 | 0.60 | 0.83 | 0.63 | 0.75 |
|  | **Median** | 0.65 | 0.70 | 0.85 | 0.79 | 0.80 |
| **Above-50%ile (n=35)** | **One Year** | 0.81 | 0.80 | 0.69 | 0.59 | 0.86 |
|  | **Two Year** | 0.81 | 0.80 | 0.63 | 0.61 | 0.86 |
|  | **Median** | 0.89 | 0.90 | 0.59 | 0.80 | 0.92 |
| **Unifocal (n=34)** | **One Year** | 0.79 | 0.69 | 0.68 | 0.72 | 0.67 |
|  | **Two Year** | 0.54 | 0.77 | 0.62 | 0.62 | 0.76 |
|  | **Median** | 0.61 | 0.79 | 0.61 | 0.75 | 0.80 |
| **Multifocal (n=37)** | **One Year** | 0.76 | 0.71 | 0.69 | 0.69 | 0.69 |
|  | **Two Year** | 0.84 | 0.80 | 0.82 | 0.80 | 0.82 |
|  | **Median** | 0.85 | 0.80 | 0.83 | 0.81 | 0.82 |

**Figure S1:**
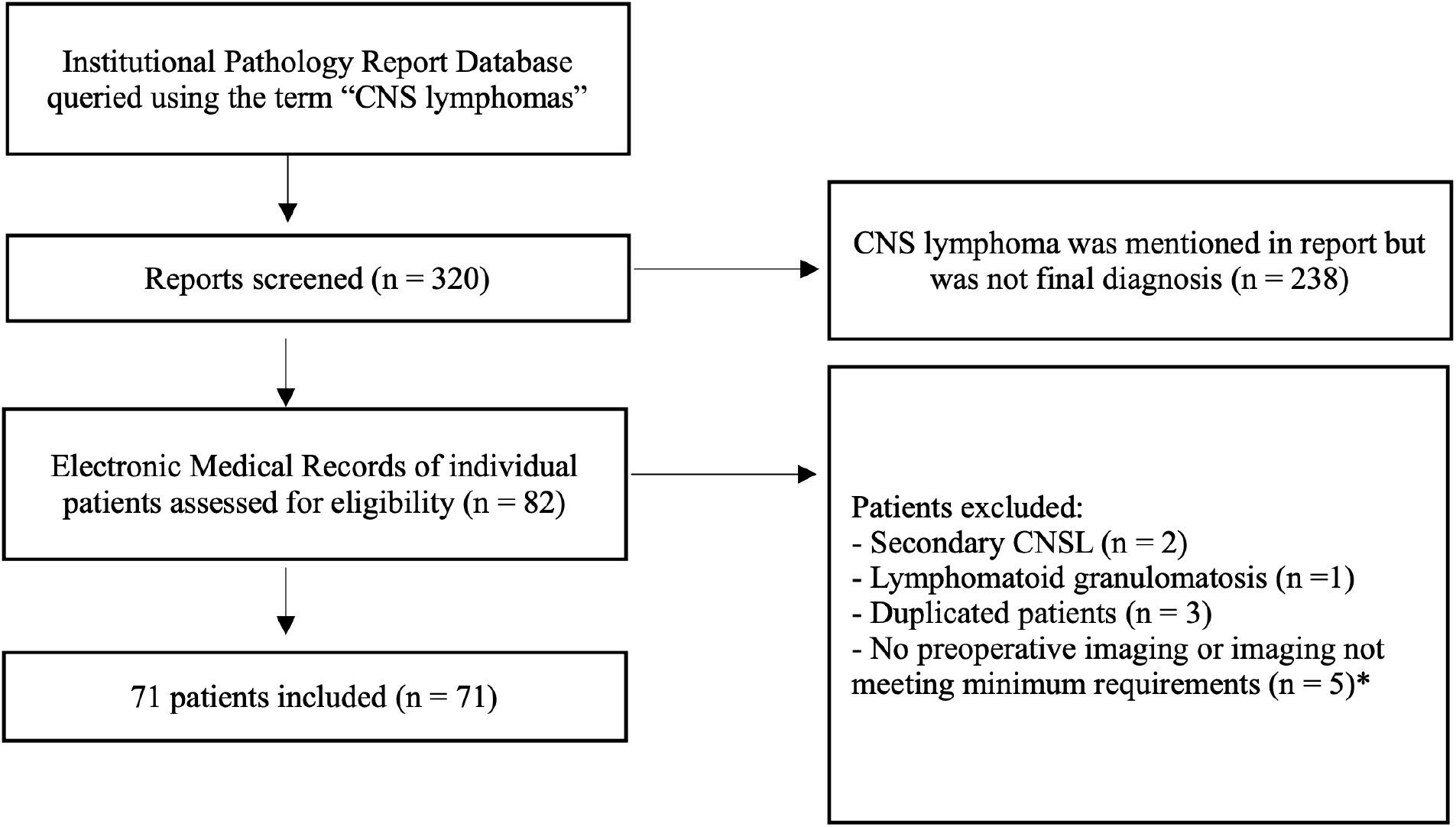
Flowchart reflection of the patient selection with inclusion and exclusion criteria.

**Figure S2:**
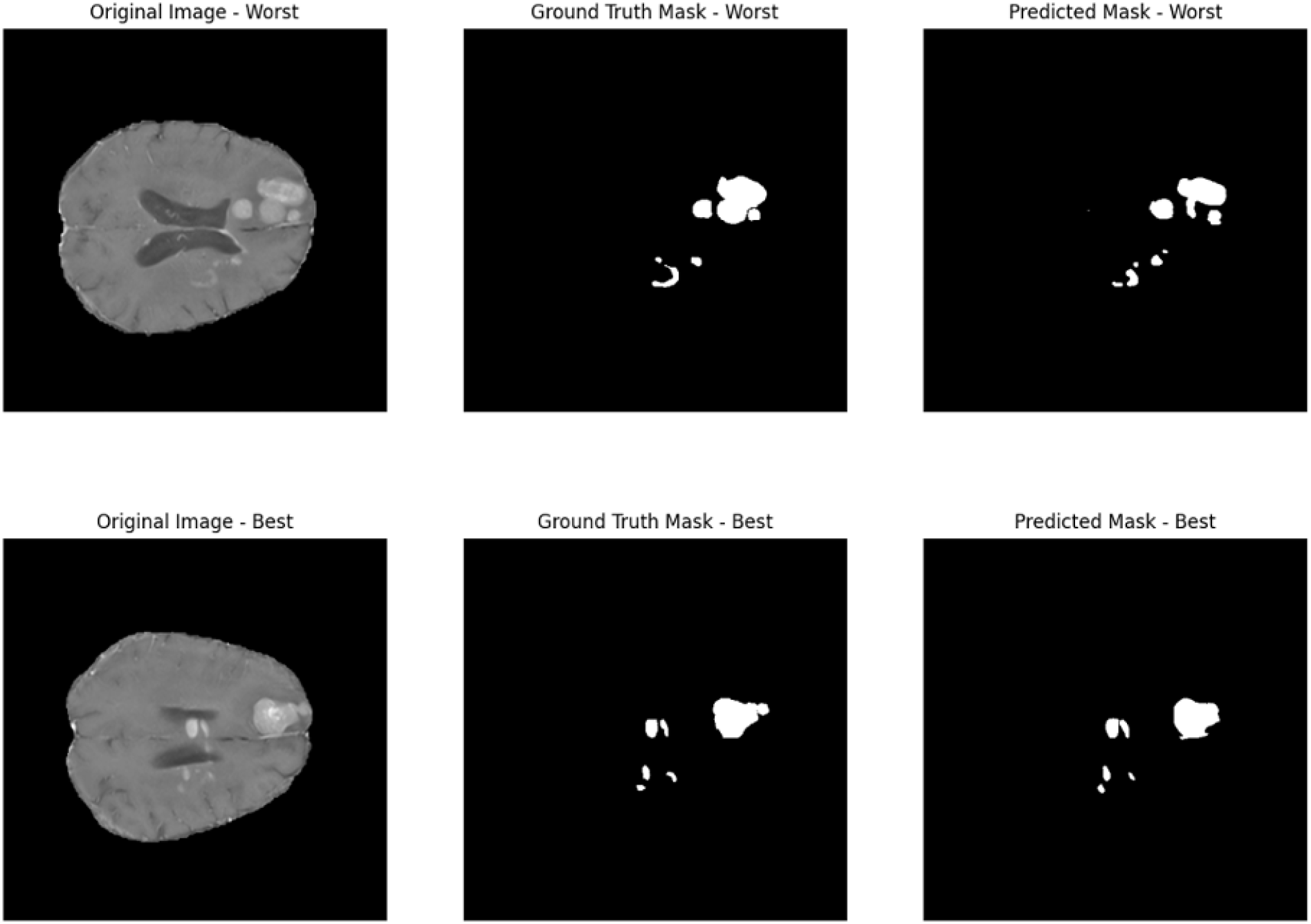
Illustration of segmentation model performance in the validation cohort, including the slice with the highest Dice coefficient (0.91) and lowest Dice coefficient (0.79) observed in our dataset.

**Figure S3:**
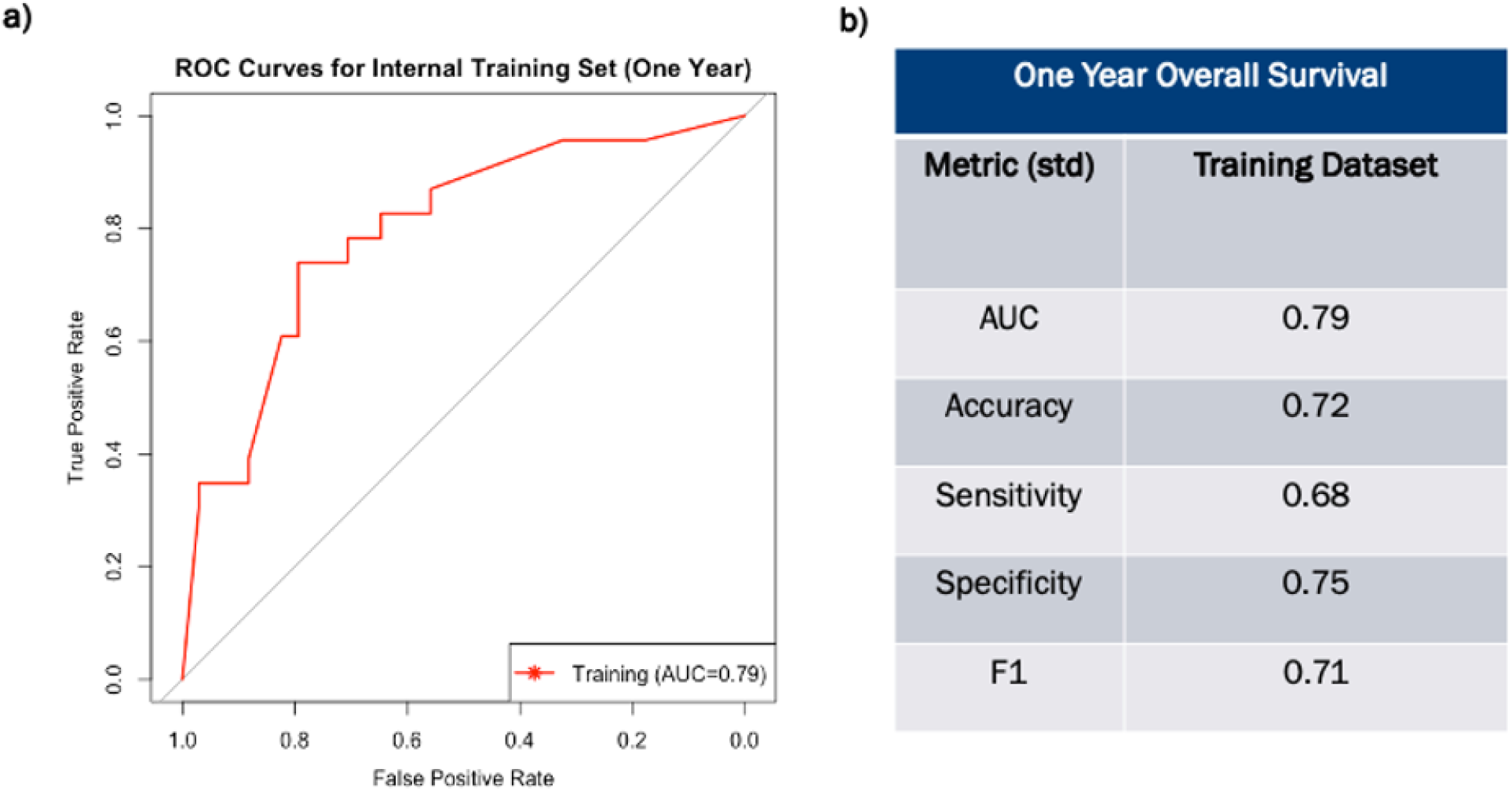
**a)** Receiver Operating Characteristic curves for one year survival model on the training cohort. Curve were generated using model logits, i.e. the raw prediction scores before binary prediction, to assess continuous prediction scores. The AUC values indicate the overall performance of each model, with higher AUC values representing better discriminative ability. **b)** Model evaluation metrics across training cohort.

**Figure S4:**
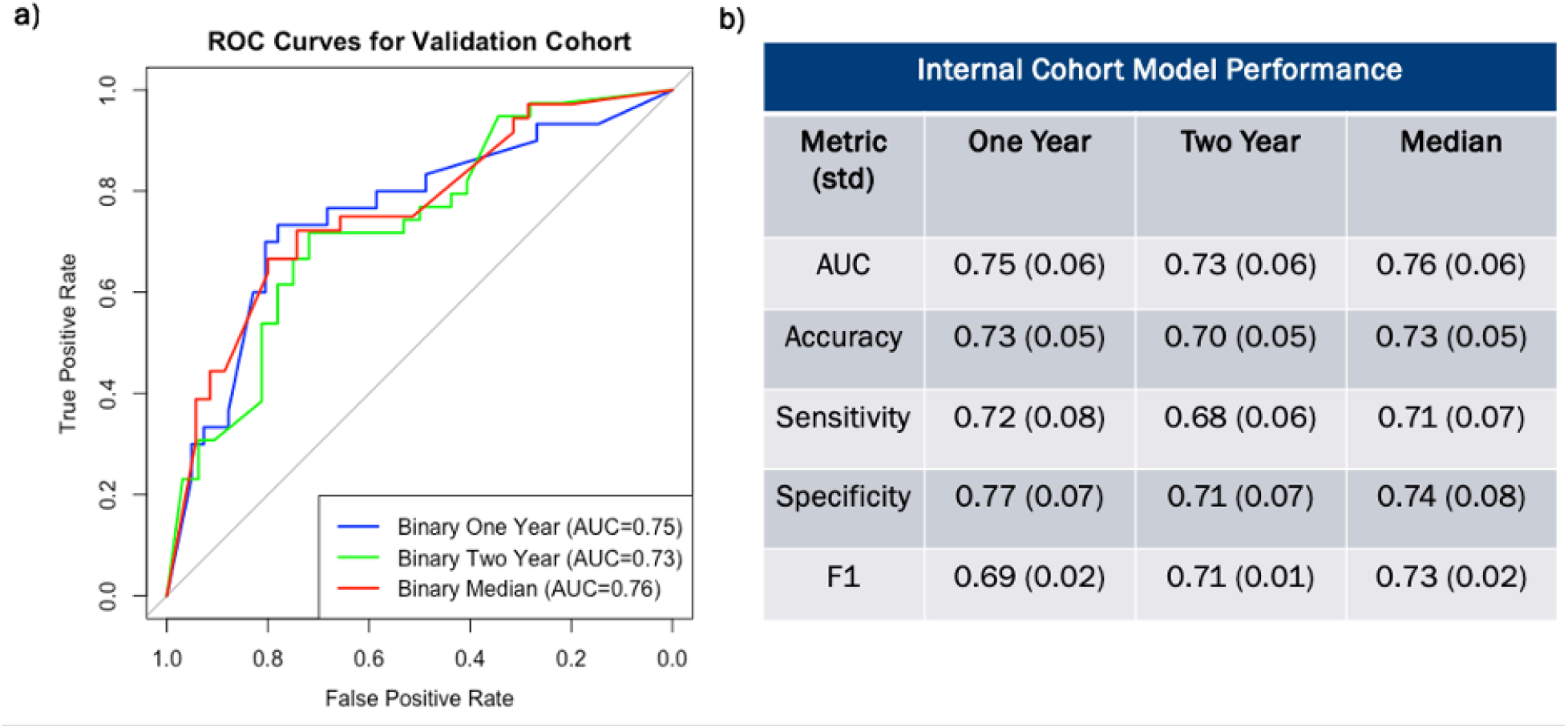
**a)** Receiver Operating Characteristic curves for three models on the internal validation cohort. Each curve was generated using model logits, i.e. the raw prediction scores before binary prediction, to assess continuous prediction scores. The AUC values indicate the overall performance of each model, with higher AUC values representing better discriminative ability. **b)** Model evaluation metric across three models.

**Figure S5:**
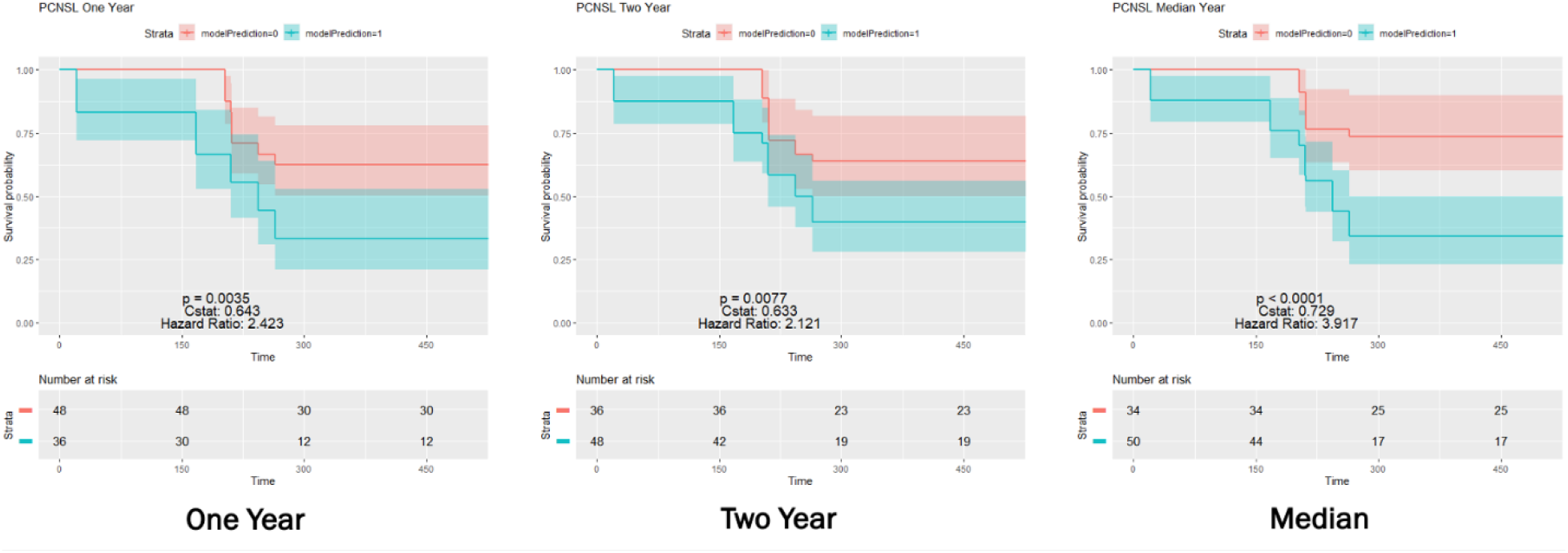
Kaplan-Meier survival curves comparing the performance of the three models across the internal cohort. Each curve represents the survival probability over time for the low-risk group (red line) and high-risk group (blue line). The log-rank test was used to assess statistical differences between the models, with p-value, C-statistic, and Hazard ratio provided in the plot.

**Figure S6:**
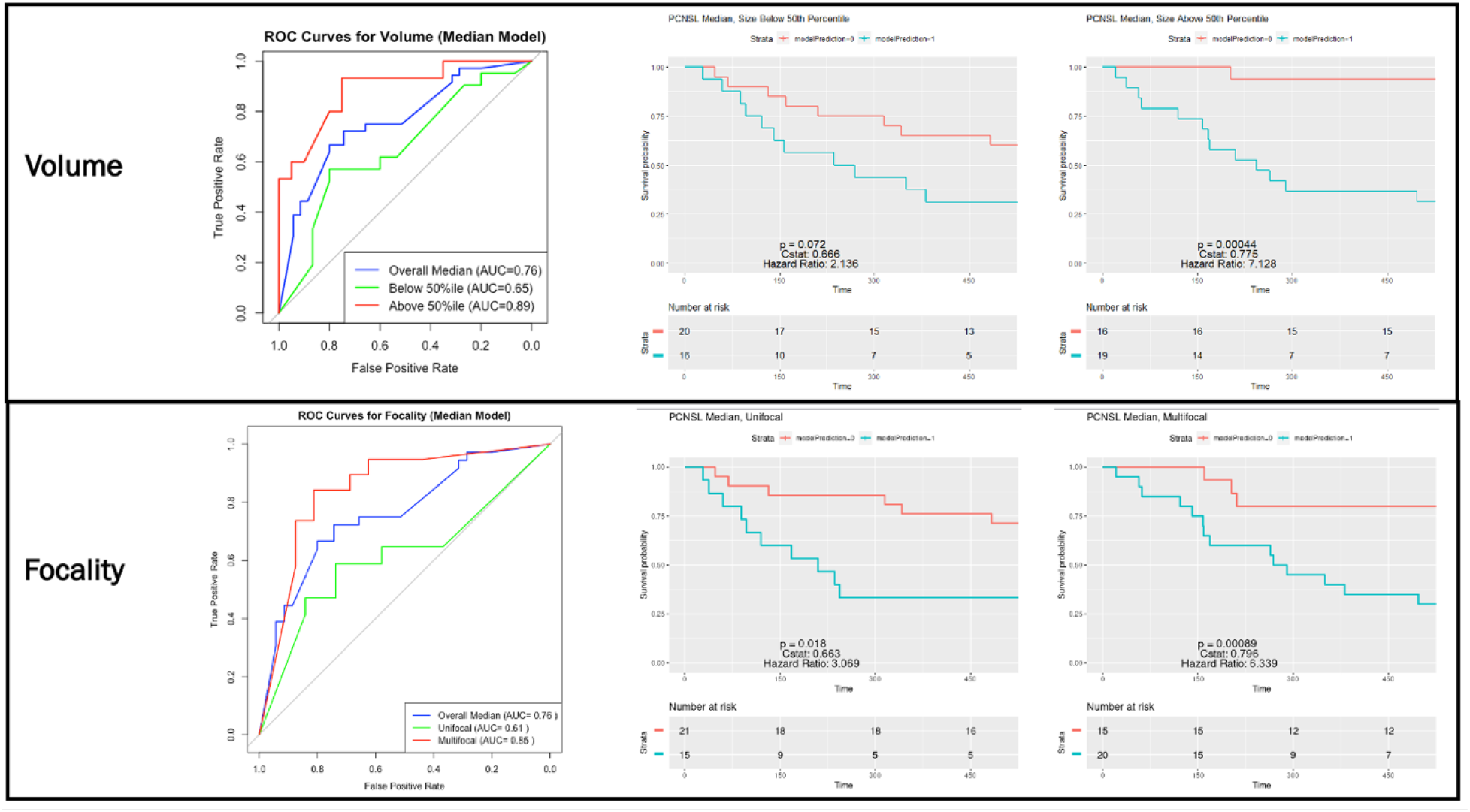
Validation of bioimaging marker across tumor size and multiplicity using median overall survival model, where the median survival of the cohort was 677 days.

**Figure S7:**
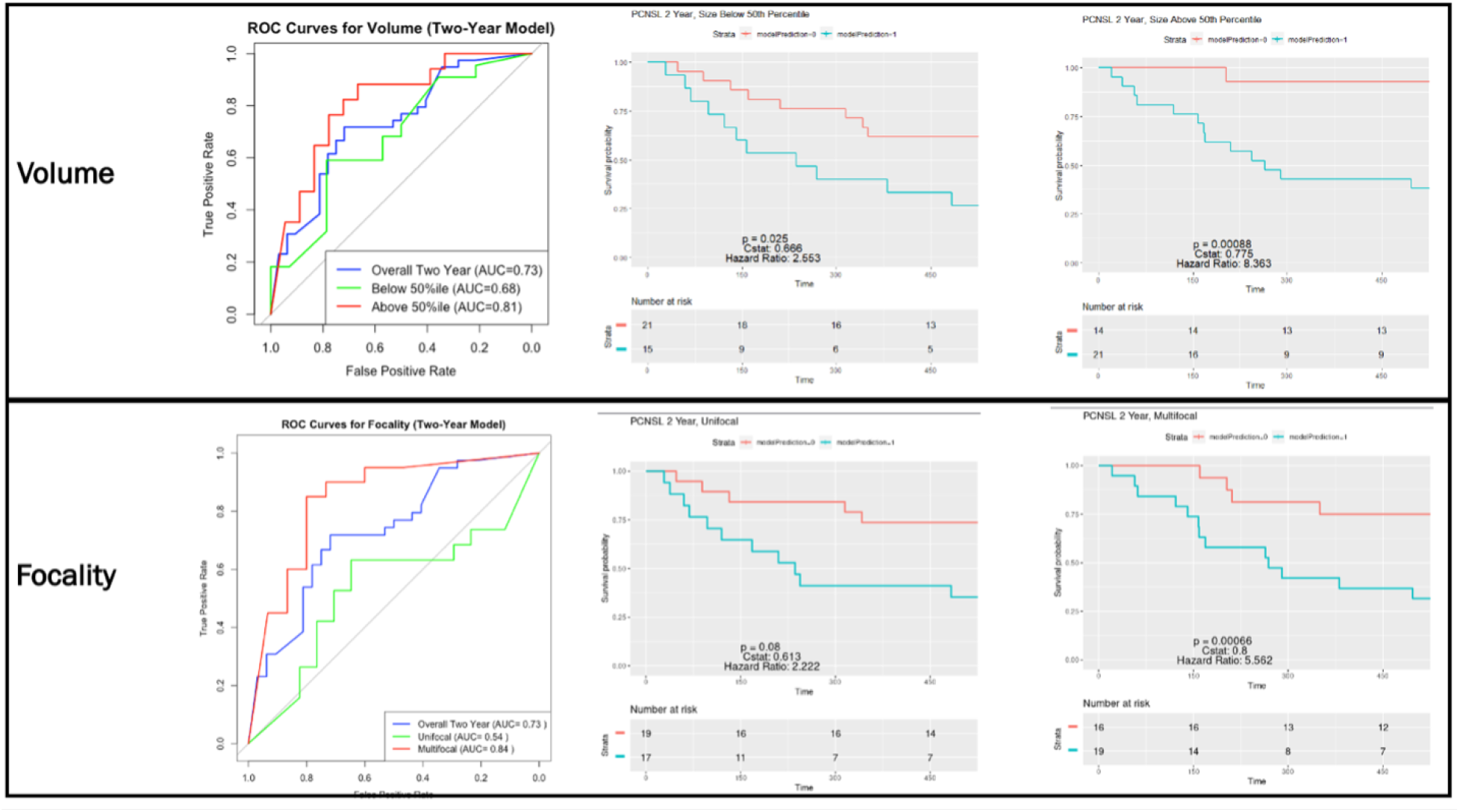
Validation of bioimaging marker across tumor size and multiplicity using two year overall survival model.

## Notes

### Competing Interest Statement

The authors have declared no competing interest.

### Funding Statement

This study did not receive any funding.

### Author Declarations

IRB of Yale University waived ethical approval for this work

### Summary of Updates

Abstract reformatting (taking out <br> tags)

